# Antibody feedback regulation of memory B cell development in SARS-CoV-2 mRNA vaccination

**DOI:** 10.1101/2022.08.05.22278483

**Authors:** Dennis Schaefer-Babajew, Zijun Wang, Frauke Muecksch, Alice Cho, Raphael Raspe, Brianna Johnson, Marie Canis, Justin DaSilva, Victor Ramos, Martina Turroja, Katrina G. Millard, Fabian Schmidt, Juan Dizon, Irina Shimelovich, Kai-Hui Yao, Thiago Y. Oliveira, Anna Gazumyan, Christian Gaebler, Paul D. Bieniasz, Theodora Hatziioannou, Marina Caskey, Michel C. Nussenzweig

## Abstract

Feedback inhibition of humoral immunity by antibodies was initially documented in guinea pigs by Theobald Smith in 1909, who showed that passive administration of excess anti-Diphtheria toxin inhibited immune responses^1^. Subsequent work documented that antibodies can enhance or inhibit immune responses depending on antibody isotype, affinity, the physical nature of the antigen, and engagement of immunoglobulin (Fc) and complement (C’) receptors^2, 3^. However, little is known about how pre-existing antibodies might influence the subsequent development of memory B cells. Here we examined the memory B cell response in individuals who received two high-affinity IgG1 anti-SARS-CoV-2 receptor binding domain (RBD)-specific monoclonal antibodies, C144-LS and C135-LS, and subsequently two doses of a SARS-CoV-2 mRNA vaccine. The two antibodies target Class 2 and 3 epitopes that dominate the initial immune response to SARS-CoV-2 infection and mRNA vaccination^4–8^. Antibody responses to the vaccine in C144-LS and C135-LS recipients produced plasma antigen binding and neutralizing titers that were fractionally lower but not statistically different to controls. In contrast, memory B cells enumerated by flow cytometry after the second vaccine dose were present in higher numbers than in controls. However, the memory B cells that developed in antibody recipients differed from controls in that they were not enriched in VH3-53, VH1-46 and VH3-66 genes and predominantly expressed low-affinity IgM antibodies that carried small numbers of somatic mutations. These antibodies showed altered RBD target specificity consistent with epitope masking, and only 1 out of 77 anti-RBD memory antibodies tested neutralized the virus. The results indicate that pre-existing high-affinity antibodies bias memory B cell selection and have a profound effect on the development of immunological memory in humans that may in part explain the shifting target profile of memory antibodies elicited by the 3^rd^ mRNA vaccine dose.

## Results

### Study design & cohorts

To examine how passive administration of monoclonal antibodies (mAbs) might influence subsequent humoral responses to vaccination, we studied a group of 18 healthy volunteers who received a single dose of the combination of two long-acting monoclonal antibodies to SARS-CoV-2 and subsequently received 2 doses of a SARS-CoV-2 mRNA vaccine (Fig 1a). The 2 antibodies, C144-LS and C135-LS, bind Class 2 and 3 epitopes on the receptor binding domain (RBD) of the SARS-CoV-2 spike (S) protein with high affinity (K_D_ =18 nM and K_D_ = 6 nM, respectively) and neutralize the virus with IC50s of 2.55 and 2.98 ng/ml, respectively^5, 8^.

**Fig. 1:**
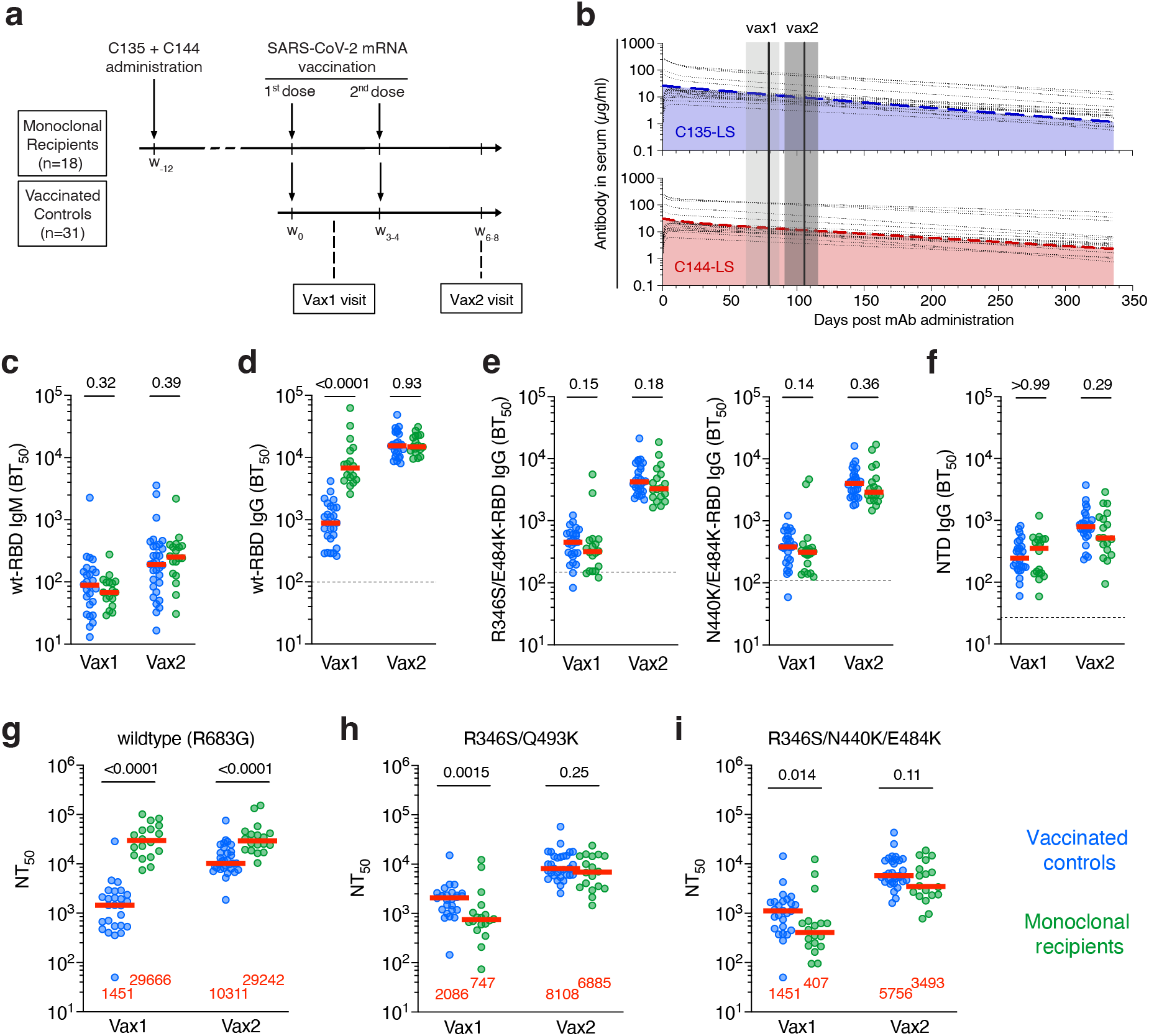
Study design and plasma antibody activity. **a,** Schematic overview of the study design with markers (w) denoting weeks relative to the time of the first vaccine dose. **b,** Serum levels of C135-LS (upper panel, in blue) and C144-LS (lower panel, in red) over time are shown. The thick colored dashed lines indicate the median serum concentrations among mAb recipients (n=18), while the thin dotted black lines represent individual participants. The two solid vertical lines indicate the median and the grey shaded areas the range of time from mAb administration. **c-f,** Half-maximal plasma binding titers (BT50) to RBD after one (vax1) and two doses (vax2) mRNA vaccinations for monoclonal antibody recipients (n=18, in green) and controls (n=26, in blue). Each dot represents one individual. Dashed horizontal lines represent the median binding activity of healthy pre-pandemic plasma samples, which served as negative controls. **c,d,** IgM (**c**) and IgG (**d**) binding titers to WT RBD. **e,** IgG binding to R346S/E484K (left panel) and N440K/E484K RBDs, see also Ext. Data Fig 1. **f,** IgG binding to the NTD. **g-i,** Plasma half-maximal neutralizing titers (NT50s) against HIV-1 pseudotyped with **g,** SARS-CoV-2 WT S. **h,** R346S/Q493K mutant S. (**i**) R346S/N440K/E484K mutant S (see also Ext. Data Fig 2). The S protein in the pseudoviruses in **g-i** contained an R683G substitution. Red horizontal bars in **c-i** and red numbers in **g-i** represent median values. Statistical significance in **c-i** was determined using the two-tailed Mann-Whitney test comparing differences between monoclonal recipients and controls for each time point independently. All experiments were performed at least in duplicate.

Between January 13 and March 3, 2021, 23 SARS-CoV-2-naïve individuals received C144-LS and C135-LS (n = 21) or placebo (n =2), in a first-in-human, phase 1 clinical trial at the Rockefeller University Hospital (NCT04700163). The antibodies were modified to extend their half-life by introducing the M428L and N343S mutations into their Fc domains^9^ (LS). Individuals received a single dose of C144-LS and C135-LS IgG1 antibodies at a 1:1 ratio, starting with 100 mg of each subcutaneously (s.c.) in the lowest- and up to 15 mg/kg intravenously (i.v.) in the highest-dose group. Participants were followed longitudinally to assess the safety and tolerability of the infused mAbs and determine their pharmacokinetic properties.

Eighteen of the 21 phase 1 study participants who had received the monoclonal antibodies elected to receive SARS-CoV-2 mRNA vaccination and volunteered to enroll in a parallel observational study assessing their immune responses to SARS-CoV-2 vaccination (Ext. Data Tables S1 and S2). The first and second vaccine doses were administered a median of 82 (range 42-110) and 103 (range 70-131) days after antibody administration (Ext. Data Tables S1 and S2). At the time of vaccination, the plasma levels of C144-LS and C135-LS were between 5 and 100 µg/ml depending on the dosing group (Fig 1b). The estimated half-lives of C144-LS and C135-LS were 69-99 days and 73-95 days, respectively.

The 18 vaccinated antibody recipients were compared to a cohort of 31 randomly selected mRNA vaccine recipients with no prior history of infection (Fig 1a and Ext. Data Tables S1, also see^10, 11^). Both groups were sampled between 13-28 (median 19) and 15-91 (median 29) days after their first and second vaccine doses, respectively. The two cohorts were relatively matched for demographic characteristics and vaccine formulation (for details see Ext. Data Tables S1 and S2) and none of the individuals included in the study seroconverted to nucleocapsid (N) at any time during observation period suggesting that they remained infection naïve.

### Plasma antibody reactivity

Plasma IgM and IgG antibody binding activity against RBD were measured by ELISA using Wuhan-Hu-1 (WT) and mutant forms of RBD (R346S/E484K and N440K/E484K) that eliminate binding by C144 and C135 but not Class 1 or 4, or some affinity-matured Class 2 or 3 antibodies (Ext. Data Fig 1, also see^11, 12^). When measured for WT RBD binding after one or two vaccine doses, the IgM titers in mAb recipients were not significantly different from controls (Fig. 1c). In contrast, IgG anti-WT-RBD titers were significantly higher in mAb recipients than in controls after one vaccine dose but equalized following the second dose (Fig. 1d, p<0.0001 and p=0.93, respectively). The initial difference was attributed to the infused monoclonals because when the same samples were tested against either R346S/E484K or N440K/E484K mutant RBDs that interfere with C144 and C135 binding, plasma IgG antibody levels in the mAb recipient samples were slightly but not significantly lower than the controls (Fig. 1e).

To determine whether the pre-existing antibodies to RBD interfered with humoral immunity to an independent domain of the SARS-CoV-2 S protein, the same plasma samples were tested for binding to the N-terminal domain (NTD). IgG titers to NTD were similar in mAb recipients and controls (Fig. 1f). We conclude that high circulating levels of C144-LS and C135-LS do not interfere with IgM anti-RBD antibody responses and have only a small effect on IgG responses. Thus, the infused antibodies do not clear the vaccine antigen or measurably interfere with its overall ability to produce an immune response^13^.

### Neutralization

To assess plasma neutralizing activity, we used HIV-1 pseudotyped with WT or mutant S proteins that carry a furin-cleavage site mutation (R683G)^14^. As expected, based on the amount of C144-LS and C135-LS in circulation, neutralizing titers against WT were significantly higher in mAb recipients than in controls at both timepoints (Fig. 1g, p<0.0001). To determine the contribution of the endogenous neutralizing response to epitopes outside of the C144-LS and C135-LS target sites, we used viruses pseudotyped with R346S/Q493K and R346S/N440K/E484K mutations that abolish the neutralizing activity of the 2 infused mAbs (Ext. Data Fig. 2). Despite the initial dominance of Class 1-2 epitopes among neutralizing antibodies elicited by mRNA vaccination^6^, the neutralizing titers of the control plasmas against the 2 mutant pseudoviruses were comparable to those against WT (Fig. 1h-i), suggesting that a significant proportion of circulating endogenous neutralizing antibodies are unaffected by the R346S/Q493K and R346S/N440K/E484K mutations. After the first vaccine dose, mAb recipients showed significantly lower neutralizing titers against the mutant pseudoviruses than controls (2.7-and 3.5-fold for R346S/Q493K and R346S/N440K/E484K; Fig. 1h and i, p=0.0015 and p=0.014, respectively). Consistent with a recent report^13^, neutralizing activity improved and was no longer significantly different from controls after the second vaccine dose (Fig 1h and i).

In conclusion, recipients of C144-LS and C135-LS had high initial levels of serum neutralizing activity due to the passively administered antibodies and they developed their own neutralizing antibodies that were not sensitive to RBD mutations in the C144/C135 target sites after mRNA vaccination.

### Memory B cells

In addition to the plasma cells that produce circulating antibodies, vaccination also elicits memory B cells that contribute to protection upon re-exposure to the pathogen. Although the feedback effects of antibodies on humoral responses have been investigated extensively beginning in 1909^1, 2^, little is known about their effects on the development of memory B cells. To investigate the effects of passive mAb administration on B cell memory responses, we used flow cytometry to enumerate and purify circulating memory B cells binding to phycoerythrin (PE) and Alexa-Fluor-647 (AF647) labeled RBDs (Ext. Data Fig 3a-c)^5^. mRNA vaccination elicited robust RBD-specific memory B cell responses in mAb recipients that were approximately 4-and 3-fold higher than in controls after the first and second vaccine doses, respectively (Fig 2a; p<0.0001 and p<0.0001, respectively). Thus, C144-LS and C135-LS administration increases the magnitude of the anti-RBD memory B cell response when compared to controls.

**Fig. 2:**
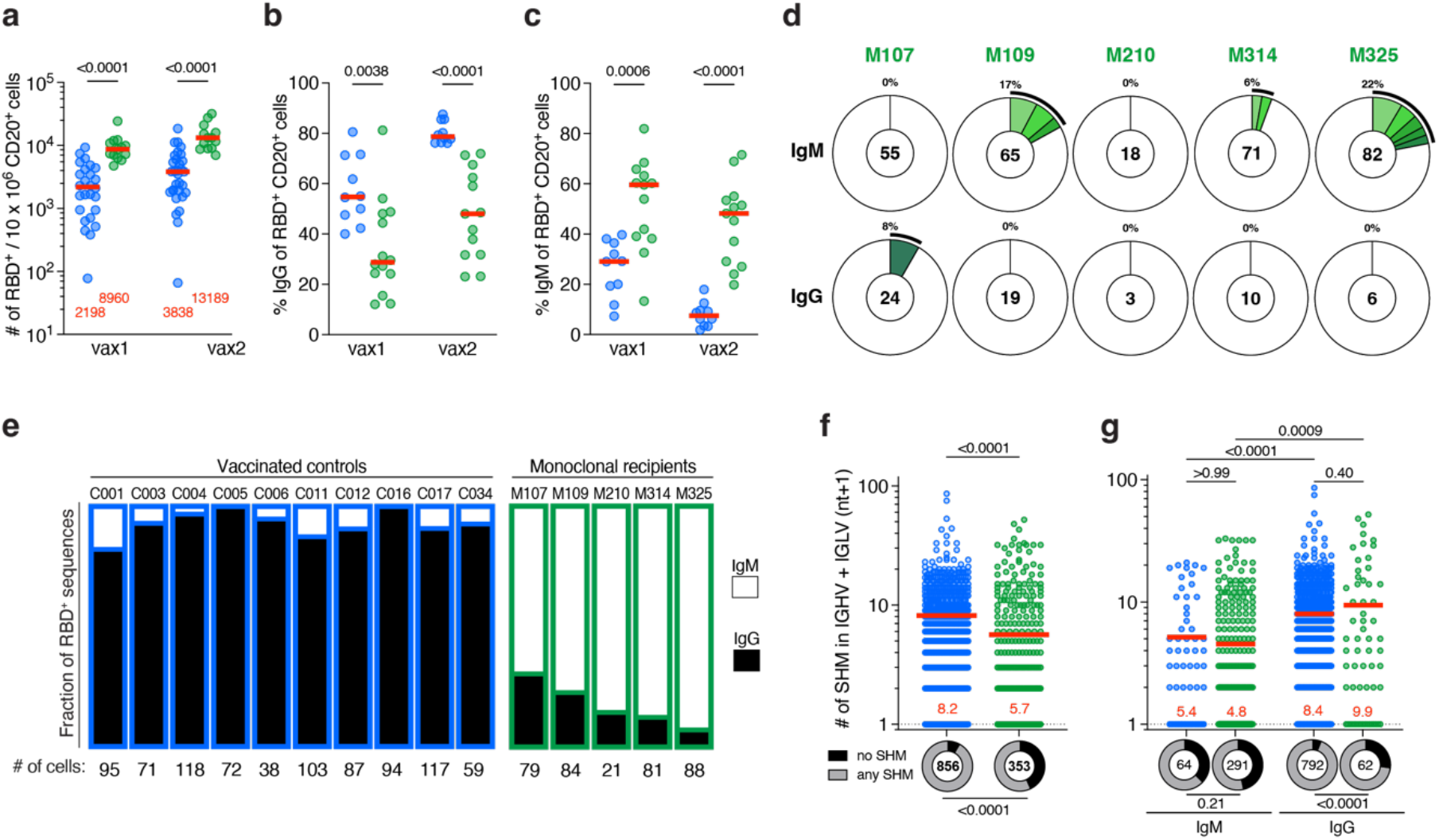
Anti-SARS-CoV-2 RBD memory B cells from vaccinated monoclonal recipients. **a-c,** Flow-cytometric enumeration and surface immunoglobulin expression of SARS-CoV-2 RBD-specific memory B cells after vax1 and vax2. mAb recipients (green, n=18 in panel **a**, n=13 in panels **b** and **c**) and controls^10, 11^ (blue, n=26 for vax1 and n=31 for vax2 in panel **a**, and n=10 in panels **b,c**). Each dot represents one individual and red horizontal bars (and numbers in panel **a**) depict median values. **a,** Number of WT RBD-specific memory B cells per 10 million CD20^+^ B cells (see also Ext. Data Fig 3a and b). **b,c,** Percentage of cells among WT RBD-binding CD20^+^ B cells that express cell surface IgG (**b**) or IgM (**c**). **d,** Pie charts show the distribution of antibody sequences derived from cells isolated from 5 vaccinated monoclonal recipients after vax2 (see also Ext. Data Fig 3d and e). The upper panel shows IgM, and the lower panel depicts IgG. The number in the inner circle indicates the number of sequences analyzed for the individual denoted above the circle. Slices colored in shades of green indicate cells that are clonally expanded (same IGHV and IGLV genes, with highly similar CDR3s) within an individual. Pie slice size is proportional to the number of clonally related sequences. The black outline and % value indicate the frequency of clonally expanded sequences detected within an individual. White pie areas indicate the proportion of sequences isolated only once. **e,** Fraction of cells harboring IgG (black) vs IgM (white) transcripts from the indicated individuals (mAb recipients outlined in green, and controls in blue). See also Ext Data Fig. 3f and ^10,11^). **f,g,** Somatic hypermutation (SHM) shown as combined heavy- and light-chain variable region nucleotide substitutions plus one (IGVH+IGVL+1), with each dot representing one sequence from mAb recipients (green) or controls (blue). Ring plots below each column show the fraction of sequences with no (IGVH+IGVL+1 = 1) vs. any (IGVH+IGVL+1 > 1) SHM, and the number in the circle indicates the number of sequences analyzed, (**f**) for all cells irrespective of isotype, (**g**) IgM and IgG analyzed independently. Red horizontal bars and numbers in **f** and **g** indicate mean values. Statistical significance was determined using the two-tailed Mann-Whitney test comparing differences between monoclonal recipients and controls for **a-c** and **f**, the Kruskal-Wallis test with subsequent Dunn’s correction for multiple comparisons was used for **g**, and Fisher’s exact test was used to compare fractions in f and **g**.

Human memory B cells represent a diverse pool of cells that can develop in germinal centers (GCs) or through an extrafollicular GC-independent pathway^15–17^. Memory B cells expressing class-switched and highly somatically mutated antibodies are primarily of germinal center origin^16, 18^. IgM-expressing memory B cells, that express antibodies that carry only small numbers of mutations typically develop by a germinal center independent pathway^19–22^. In control individuals, IgG-expressing RBD-specific memory cells comprised the majority of the memory B cell pool at both time points assayed with many fewer such cells found in mAb recipients (p=0.0038 and p<0.0001, Fig 2b). Consistent with the relative decrease in IgG^+^ memory B cells, nearly two thirds (65%) of the RBD-specific memory B cells from mAb recipients were cell surface IgM^+^ after the first vaccine dose and this decreased only slightly to 54% after the second vaccine dose (Fig 2c). We conclude that pre-existing high-affinity anti-RBD antibodies alter the immune response to SARS-CoV-2 mRNA vaccination to favor the development of IgM-expressing memory B cells.

### Memory B cell Antibodies

To gain further insight into the effects of pre-existing antibodies on the human memory response to SARS-CoV-2 mRNA vaccination, we purified RBD-specific memory B cells from 5 mAb recipients after the second vaccine dose. A total of 353 and 856 paired antibody sequences from mAb recipients and previously characterized controls were examined, respectively (Fig 2d, Ext. Data Fig. 3d-f and Ext. Data Table 3, also see^10^). IgM transcripts accounted for 70-94% of sequences recovered from mAb recipients with an average of 9% belonging to expanded clones (Fig 2d, upper panel and Fig 2e). In contrast, IgG transcripts accounted for >90% of the immunoglobulin sequences isolated from controls (Fig 2e and Ext. Data Fig 3f). IgM memory cells originating from the extrafollicular non-GC pathway are generally less somatically mutated than IgG memory cells because they undergo fewer divisions^16, 22^. Consistent with this idea, and the reversed ratio of IgM:IgG memory B cells in mAb recipients, the antibodies obtained from these individuals showed significantly fewer somatic mutations than controls (Fig. 2f, p<0.0001). However, when comparing IgM or IgG cells independently, the average mutational burden was not significantly different between mAb recipients and controls (Fig 2g, p>0.99 and p=0.40 for IgM and IgG, respectively). Thus, IgM-and IgG-expressing B cells in vaccinated individuals who had received C144-LS and C135-LS carry normal numbers of somatic mutations, but the relative ratio of the two memory cell types is reversed, which accounts for the overall lower level of mutation in the memory compartment. Finally, in contrast to controls there was no enrichment for VH3-53, VH1-69, VH1-46 and VH3-66 heavy chains, which often target Class 1 and 2 epitopes. Instead, there was relative enrichment for VH3-9, VH5-51, VH4-39 and VH1-8 genes (Ext Data Fig. 4a). In summary, the data suggest that pre-existing antibodies can alter the cellular and molecular composition of the RBD-specific MBC compartment that develops in response to mRNA vaccination.

To examine the binding and neutralizing activity of the memory antibodies elicited by mRNA vaccination in C144-LS and C135-LS recipients, we produced 178 representative monoclonals obtained from 5 individuals as IgGs and tested them for binding to the WT SARS-CoV-2 RBD by ELISA (Fig. 3a-c and Ext. Data Table 4). In contrast to controls, where over 95% of the memory antibodies bound strongly to RBD, monoclonal antibodies isolated from volunteers that received C144-LS and C135-LS showed diverse levels of binding activity. Approximately one quarter (24%) of the antibodies displayed relatively poor binding with ELISA half-maximal effective concentrations (EC50s) that were only slightly above our limit of detection, and a little over one third (38%) showed no detectable binding above background (Fig. 3a-b). Accordingly, the median (EC50) of antibodies isolated from mAb recipients was significantly higher than in controls (Fig. 3b, p<0.0001). Notably, this difference remained significant when the monoclonals isolated from IgM and IgG memory cells were analyzed independently (Fig 3c, p=0.0005 and p<0.0001, respectively).

**Fig. 3:**
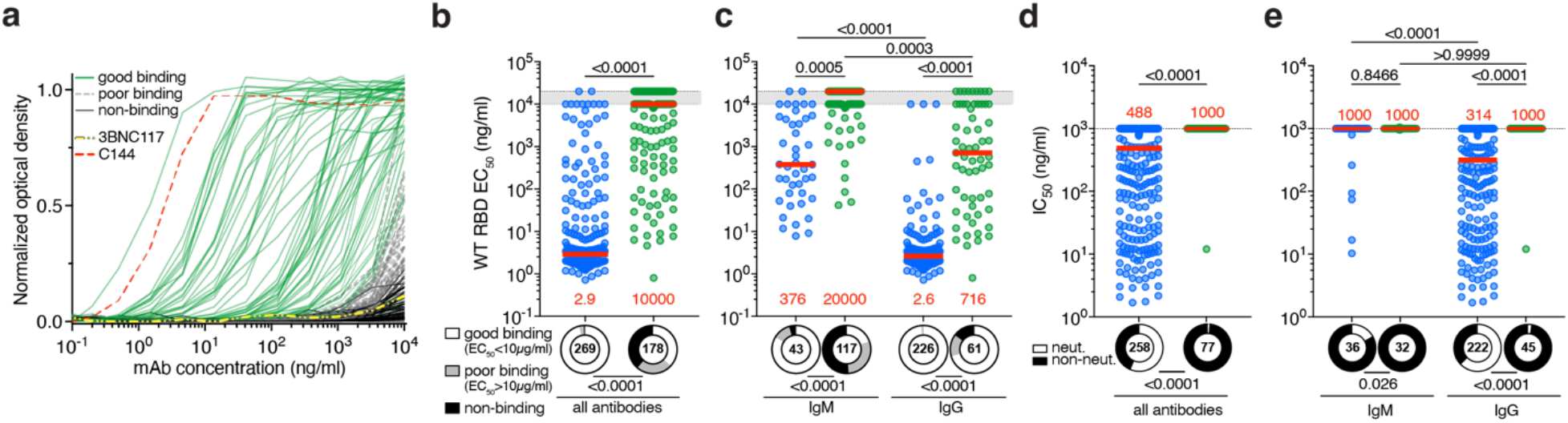
Functional characterization of anti-SARS-CoV-2 RBD memory antibodies from vaccinated mAb recipients. **a-c,** Monoclonal antibody binding to WT RBD. **a,** Graph shows ELISA binding monoclonal antibodies derived from mAb recipients after serial dilution. Each curve represents one antibody. Green curves show EC50s <10 µg/ml, grey dashed lines EC50s >10 µg/ml, solid black lines are antibodies that were below or equal to a negative control anti-HIV1 antibody 3BNC117 (thick, white-dashed line). C144 (thick, red-dashed line) is positive control. **b** Summary of EC50s derived from (**a**) mAb recipients in green, and controls in blue for all antibodies irrespective of isotype. **c,** as in (**b)** but IgM and IgG analyzed independently. Grey shaded area between horizontal dotted lines indicates antibodies with EC50s >10 µg/ml (poor binding) and non-binding antibodies arbitrarily grouped at 10 and 20 µg/ml, respectively. Ring plots summarize the fraction of all antibodies tested for the respective groups (encircled number). **d,** Plots show IC50s for all monoclonal antibodies isolated from vaccinated mAb recipients (green) or controls (blue). Ring plots illustrate the fraction of non-neutralizing (IC50 > 1000 ng/ml) antibodies (black slices) among all antibodies tested for the respective group (encircled number). **e,** as in (**d**) but IgM and IgG antibodies analyzed independently. For panels **b-e**, red horizontal bars and numbers represent median values. Statistical significance was determined using the two-tailed Mann-Whitney test for **b** and **d**, whereas the Kruskal-Wallis test with subsequent Dunn’s correction for multiple comparisons was used for **c** and **e**. To compare fractions from ring plots, the Chi-squared contingency statistic was used in **b** and **c**, and Fisher’s exact test for **d** and **e**. All experiments were performed at least in duplicate.

Memory antibodies obtained from C144-LS and C135-LS recipients that bound to WT SARS-CoV-2 RBD with EC50s <10 µg/ml were tested for neutralizing activity against viruses pseudotyped with WT spike. Whereas almost two thirds (63%) of the IgG and 17% of the IgM antibodies isolated from controls showed measurable neutralizing activity, only 1 out of 45 IgG and none of the 32 IgM antibodies obtained from C144-LS and C135-LS recipients neutralized SARS-CoV-2 (Fig. 3d-e). Thus, the antibodies isolated from the RBD-specific memory B cell compartment of vaccinated mAb recipients show significantly less neutralizing activity than controls.

To examine the affinity of the antibodies, we performed biolayer interferometry experiments (BLI) in which monoclonal antibodies were immobilized on the biosensor chip and exposed to WT RBD monomers^5^ (Fig. 4a and b). In contrast to controls, where 96% of the antibodies tested displayed measurable affinities, only two thirds (67%) of the antibodies derived from mAb recipients did so (Fig. 4b, d and f, p<0.0001). When all antibodies were considered together, the median affinity (Kd) differed by nearly one order of magnitude between mAb recipients and controls (Fig. 4f, p<0.0001). Moreover, this difference remained significant when IgM and IgG monoclonals were considered independently (Fig 4g, p =0.0058 and p<0.0001, respectively), indicating that the lower affinities observed in the memory compartment of mAb recipients cannot solely be explained by the preponderance of IgM.

**Fig. 4:**
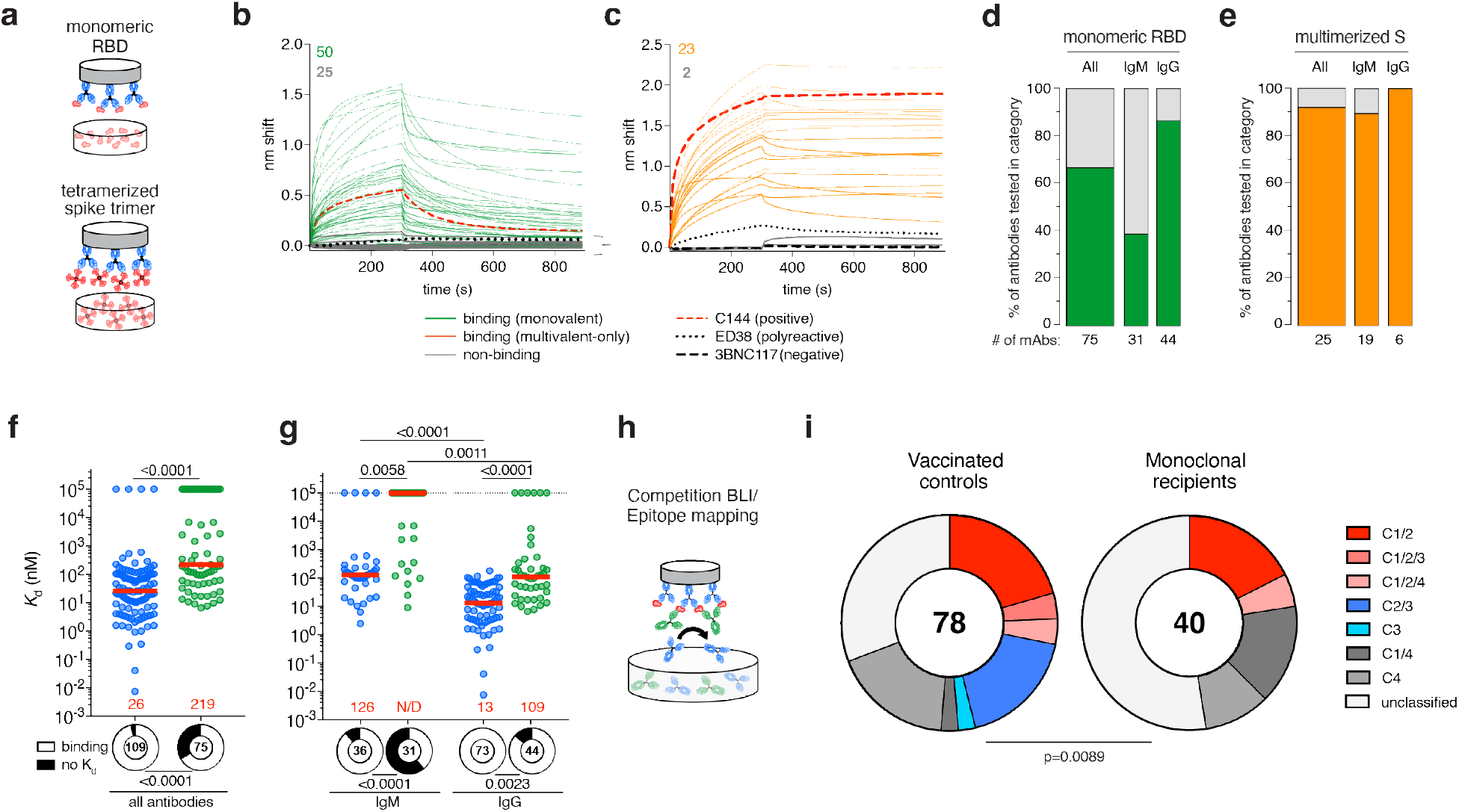
Affinities and epitope distribution of anti-SARS-CoV-2 RBD memory antibodies from vaccinated mAb recipients. **a-g,** Monoclonal antibody binding to monomeric and multimerized antigen by BLI. **a,** Schematic representation for monomeric binding measurements where IgG is immobilized on the biosensor chip and subsequently exposed to monomeric RBD (upper panel), and multimeric binding using 6P-stabilized WT SARS-CoV-2 S protein trimers that had been tetramerized using streptavidin (lower panel). **b,** Graphs show BLI traces obtained under monovalent conditions. Each curve represents one antibody. Colored solid lines denote binding above background represented by polyreactive antibody ED38^28^ (dotted black line) and anti-HIV-1 antibody 3BNC117 (dashed black line). Grey lines show non-binding antibodies. C144 (thick, red-dashed line) is a positive control. Colored and grey numbers in upper left of each panel indicate the number of binding and non-binding antibodies, respectively. **c**, As in (**b**) for antibodies that showed no measurable binding in (**b**) and were subsequently tested for binding under polyvalent conditions. **d,** Bar charts show the percentage of binding antibodies under monovalent conditions for all antibodies and by isotype. Values below bars indicate the number of antibodies tested. **e,** as in (**d**) for antibodies shown in (**c**). **f,** Graphs show affinity constants (K_d_) derived under monomeric binding conditions (**b)** for mAb recipients (green) and controls (blue) irrespective of isotype. Ring plots illustrate the fraction of antibodies tested for the respective group (encircled number) that measurably bound to monomeric RBD (“binding”, in white) and those for which a K_d_ value could not be established (“no K_d_”, black). Red horizontal bars and numbers represent median values (N/D, not determined). **g**, as in (**f**) analyzed independently for IgM and IgG. **h**, Schematic representation of BLI competition experiment in which a capture antibody of known epitope-specificity (class-reference antibody) is bound to the biosensor chip and exposed to antigen. In a second step, the antibody of interested is added to the chip. **i**, Pie charts show the distribution of epitopes targeted. The number in the center is the number of antibodies tested. Slices colored in shades of red and blue represent Class 1, 2 and 3 or combined epitopes, shades of grey represent Class 4-containing epitopes or epitopes that could not be classified by this method. Statistical significance was determined using the two-tailed Mann-Whitney test for **f**, and the Kruskal-Wallis test with subsequent Dunn’s correction for multiple comparisons for **g**. To compare categories and distributions from ring plots, Fisher’s exact test was used for **f** and **g**, and the Chi-squared contingency statistic was used for panel **i**.

C144-LS and C135-LS have the potential to form immune complexes with the vaccine antigen *in vivo* and present it as a multimer that could increase the apparent affinity of a B cell for the multimerized antigen by avidity effects. To determine whether memory cell-derived antibodies from mAb recipients with no apparent affinity to monomeric antigen would show binding under higher valency conditions, we exposed the immobilized monoclonals to biotin-streptavidin tetramerized trimers of S (Fig. 4a, c and e). Of the 25 antibodies with no apparent monomeric binding tested, 23 (92%) bound to multimerized S (Fig. 4c and e). We conclude that most of the anti-RBD antibodies isolated from mAb recipients that failed to show detectable binding to RBD monomers bind to multimerized antigen.

To examine the epitopes targeted by the vaccine-elicited anti-RBD antibodies produced by memory B cells of C144-LS and C135-LS recipients, we performed BLI experiments in which a pre-formed antibody-RBD complex composed of one of 4 structurally characterized antibodies^8, 23, 24^ was exposed to a second monoclonal targeting an unknown epitope (Fig 4h and Ext. Data Fig. 5). 49% of the anti-RBD memory antibodies obtained from vaccinated controls target Class 1, 2 or 3 epitopes or combinations thereof (Fig. 4i, Ext. Data Fig 5a-e, and ^10, 11^). In contrast, only 20% of the memory antibodies obtained from mAb recipients targeted Class 1 or 2 epitopes, and none were Class 3-specific. Instead, we found that 78% of these antibodies targeted either Class 4-containing epitopes or epitopes that could not be classified by our method (Fig. 4i and Ext. Data Fig 5a-e). Thus, there was a significant shift in the distribution of epitopes targeted by memory antibodies isolated from mAb recipients compared to controls (Fig. 4i, p=0.0089). In conclusion, C144-LS and C135-LS alter the development of memory B cells expressing anti-RBD antibodies and their epitope target preference.

## Discussion

Our experiments show that pre-existing antibodies alter the development of memory B cells in response to SARS-CoV-2 mRNA vaccination in humans. Consistent with a recent report, C144-LS and C-135-LS did not significantly interfere with the development of circulating antibodies that bind to epitopes outside of the target sites of the two monoclonal antibodies^13^. And while we found that neutralizing responses were reduced in mAb recipients after one dose, this difference was no longer significant after two doses of mRNA vaccination. In contrast, anti-RBD specific memory B cell development was profoundly altered. Memory cells expressing IgG antibodies specific to Class 1, 2 or 3 epitopes normally dominate the anti-RBD response after 2 doses of mRNA vaccination^4–8^. In contrast, mAb recipients develop increased numbers of anti-RBD memory cells that express low-affinity IgM antibodies with altered epitope specificity.

Beginning with experiments on anti-Diptheria toxin antibodies in the early part of the 20^th^ century, extensive work in experimental animals showed that passive transfer of polyclonal immune serum or monoclonal antibodies can alter subsequent humoral immune responses in an epitope-specific manner^1, 2^. However, the effect of pre-existing antibodies on the development of memory B cells is not well understood. C144 and C135 bind to Class 2 and 3 epitopes on the RBD and interfere with the development of memory responses to the normally dominant classes of antibodies by what appears to be epitope masking. Epitope masking may also explain the shift in memory B cell specificity away from Class 1 and 2 after the 3^rd^ dose of the SARS-CoV-2 mRNA vaccines that increases the breadth of the neutralizing response^11^. In chronic infections, such as HIV-1, masking could account for the shift from initial anti-gp41 responses to other parts of the trimer^25, 26^ and it could interfere with the development of broad neutralizing responses to HIV-1 or influenza by shifting memory responses away from conserved epitopes^27^.

Memory B cells can develop by 2 different pathways^15, 16^. Class-switched memory B cells that carry relatively higher-affinity antibodies with large numbers of somatic mutations develop in germinal centers (GCs). In contrast, IgM-expressing memory B cells that carry lower-affinity antibodies and only small numbers of mutations develop by a GC-independent pathway^16, 17^. Passive transfer of C144-LS and C135-LS may favor the GC-independent pathway by creating immune complexes that contain multiple copies of the antigen in a form that increases apparent affinity by avidity effects, thereby enabling recruitment of B cells with very low affinity receptors into the immune response. A reduction in selection stringency may also explain the greater total number of RBD-binding cells found in mAb recipients. In addition, pre-existing high affinity antibodies may alter secondary GC responses by favoring entry of cells expressing lower-affinity antibodies that target a further diversified group of epitopes. In contrast, low-affinity polyclonal antibodies emerging after vaccine priming may enhance vaccine booster responses by mechanisms that remain to be fully elucidated^2, 3^.

In conclusion, the development of memory in response to vaccination is influenced by pre-existing antibodies that can alter the antibody target profile, affinity and isotype of the responding cells. Diversification of the antibody response by this mechanism may help increase the breadth of vaccines like the SARS-CoV-2 vaccine but interfere with the development of breadth in others, like HIV-1 or influenza, by diverting immunity away from broadly neutralizing to strain-specific epitopes.

## Methods

### Study participants

Participants in the “monoclonal antibody recipient” group were healthy SARS-CoV-2-naïve volunteers who were enrolled in a first-in-human phase 1 study at the Rockefeller University Hospital in New York and received single doses of two anti-SARS-CoV-2 RBD monoclonal antibodies (mAbs), C144-LS and C135-LS, between January 13 and March 3, 2021 (NCT04700163). The phase 1 clinical trial had a dose escalation design and evaluated the safety and tolerability, and pharmacokinetics of the two mAbs. The mAb cocktail (1:1 ratio of C144-LS and C135-LS) was administered to 21 out of the 23 enrolled individuals (n=2 receiving placebo), allowing for multiple interim safety analyses. The mAbs were administered at 100 or 200 mg each, subcutaneously, or at 1, 5 or 15 mg/kg each, intravenously (see Ext. Data. Table S2). Eligible participants for the phase 1 study were healthy adults with no history of SARS-CoV-2 infection or vaccination, or prior receipt of any SARS-CoV-2 therapeutics, including other monoclonal antibodies or convalescent plasma. Further details on inclusion and exclusion criteria, study design and endpoints of the phase 1 study can be found on clinicaltrials.gov (NCT04700163). Of the 21 individuals who received the mAbs in the phase 1 study, 18 co-enrolled in a parallel observational study to assess their immune responses to subsequent SARS-CoV-2 mRNA vaccination. One individual chose to receive the Janssen (Ad26.COV2.S) vaccine, and another individual (a placebo recipient) displayed Nucleocapsid (N) titer changes prior to enrollment in the observational study that were compatible with a recent SARS-CoV-2 infection, making them ineligible for inclusion in this study. The remaining phase 1 study participant chose not to enroll in the parallel study of immune responses. The 18 mAb-recipients included in this observational study received either the Moderna (*Spikevax,* mRNA-1273) or Pfizer-BioNTech (*Comirnaty,* BNT162b2) mRNA vaccines against the wildtype (Wuhan-Hu-1) strain of the severe acute respiratory syndrome coronavirus 2 (SARS-CoV-2). Participants in the “vaccinated control” group were healthy SARS-CoV-2-naïve volunteers who had received two doses of one of the two currently approved SARS-CoV-2 mRNA vaccines, Moderna (*Spikevax,* mRNA-1273) or Pfizer-BioNTech (*Comirnaty,* BNT162b2) mRNA vaccines. These control individuals had been recruited to the Rockefeller University Hospital for serial blood donations to longitudinally assess their immune responses to SARS-CoV-2 mRNA vaccination^10, 11^. We previously reported the findings obtained from this group and refer to Cho et al.^10^ and Muecksch et al.^11^ for further details on participant recruitment, inclusion and exclusion criteria, and demographic characteristics (also see Ext. Data Tables S1 and S2). At each sample collection visit, participants of either group presented to the Rockefeller University Hospital for blood sample collection and were asked to provide details of their vaccination regimen, possible side effects, comorbidities, and possible COVID-19 history. Vaccinations were administered outside of the study, at the discretion of the individual and their health care provider consistent with existing guidelines and, as such, not influenced by their participation in our study. Baseline and longitudinal plasma samples were tested for binding activity toward the nucleocapsid protein (N) (Sino Biological, 40588-V08B) of SARS-CoV-2. Absence of seroconversion toward N during the study interval was used to exclude SARS-CoV-2 infection, in addition to participants’ reported history. Clinical data collection and management were carried out using the software iRIS by iMedRIS (v. 11.02). All participants provided written informed consent before participation in the study, which was conducted in accordance with Good Clinical Practice. The study was performed in compliance with all relevant ethical regulations, and the clinical protocols (CGA-1015 and DRO-1006) for studies with human participants were approved by the Institutional Review Board of the Rockefeller University. For detailed participant characteristics see Supplementary Tables 1 and 2 and ^10, 11^

### Blood samples processing and storage

Peripheral Blood Mononuclear Cells (PBMCs) obtained from samples collected at Rockefeller University were purified as previously reported by gradient centrifugation and stored in liquid nitrogen in the presence of Fetal Calf Serum (FCS) and Dimethylsulfoxide (DMSO)^5^. Heparinized plasma and serum samples were aliquoted and stored at −20°C or less. Prior to experiments, aliquots of plasma samples were heat-inactivated (56°C for 1 hour) and then stored at 4°C.

### Pharmacokinetics of C144-LS and C135-LS

To evaluate the pharmacokinetic (PK) properties of the passively administered antibodies, C144-LS and C135-LS, their serum antibody levels were measured on the day of antibody administration (day 0) at 1, 3,6, 9 and 12 hours after infusion, and on days 1, 3, 7, 14, 21, 28, 56, 84, 126, 168, 252, and 336. C144-LS and C135-LS levels in serum were measured by mass-spectrometry (MS/MS). Briefly, analytes were isolated from serum samples through immunocapture using streptavidin beads and biotinylated RBD protein. The isolated proteins were denatured with dithiothreitol, alkylated with iodoacetamide, and digested with trypsin. The final extract was analyzed via high-performance liquid chromatography (HPLC) with column-switching and MS/MS detection using positive ion electrospray. A linear, 1/concentration² weighted, least-squares regression algorithm was used for quantification. Noncompartmental analysis (NCA) was used to estimate PK parameters from measured serum levels of C144-LS and C135-LS. Phoenix WinNonlin® (v8.2) was used for the NCA. Actual sample time post administration of each mAb was used for the estimation of serum PK parameters instead of nominal time. Half-life estimates were similar between administration routes for both C144-LS and C135-LS, indicating a half-life of 2-3 months for both mAbs by either administration route (C144-LS: 68.9 to 99.3 days for s.c. groups and 86.9 to 92.3 days for i.v. groups; C135-LS: 72.7 to 77.9 days for s.c. groups and 70.5 to 94.7 days for i.v. groups). Visualization of the PK data was performed in GraphPad Prism, using the three-phase decay model.

### ELISAs

Enzyme-Linked Immunosorbent Assays (ELISAs)^29, 30^ to evaluate antibodies binding to SARS-CoV-2 Wuhan-Hu-1 RBD, NTD or S were performed by coating of high-binding 96-half-well plates (Corning 3690) with 50 μl per well of a 1μg/ml protein solution in Phosphate-buffered Saline (PBS) overnight at 4°C. Plates were washed 6 times with washing buffer (1× PBS with 0.05% Tween-20 (Sigma-Aldrich)) and incubated with 170 μl per well blocking buffer (1× PBS with 2% BSA and 0.05% Tween-20 (Sigma)) for 1 hour at room temperature. Immediately after blocking, monoclonal antibodies or plasma samples were added in PBS and incubated for 1 hour at room temperature. Plasma samples were assayed at a 1:66 starting dilution and serially diluted by either three-or fourfold. Monoclonal antibodies were tested at 10 μg/ml starting concentration and 11 additional threefold serial dilutions. Plates were washed 6 times with washing buffer and then incubated with anti-human IgG or IgM secondary antibody conjugated to horseradish peroxidase (HRP) (Jackson Immuno Research 109-036-088 and 109-035-129) in blocking buffer at a 1:5,000 dilution (IgM and IgG). Plates were developed by addition of the HRP substrate, 3,3’,5,5’-Tetramethylbenzidine (TMB) (ThermoFisher) for 6 minutes. The developing reaction was stopped by adding 50 μl of 1 M H_2_SO_4_ and absorbance was measured at 450 nm with an ELISA microplate reader (FluoStar Omega, BMG Labtech) with Omega and Omega MARS software for analysis. Normalizer control samples were included on each plate. For plasma samples and monoclonal antibodies half-maximal binding titers (BT50s) and half-maximal effective concentrations (EC50s), respectively, were calculated using four-parameter nonlinear regression (GraphPad Prism V9.3, with the following settings: [Agonist] vs. response -- Variable slope (four parameters), bottom=0, Hillslope>0, Top=plate/experiment-specific upper plateau of the normalizer control antibody/plasma reaching saturation for at least 3-consecutive dilution steps. The curve-fit was constrained to an upper limit that corresponds to the maximal optical density achieved by the known normalizer control to limit inter-plate-/experiment variability (batch effects).

Pre-pandemic plasma samples from healthy donors and isotype control monoclonal antibodies served as negative controls as indicated and were used for validation (for more details see ^5^). All reported EC50 and BT50 values are the average of at least 2 independent experiments.

### Proteins

The mammalian expression vector encoding the Receptor Binding-Domain (RBD) of SARS-CoV-2 (GenBank MN985325.1; Spike (S) protein residues 319-539) was previously described^23^. Plasmids encoding the R346S/E484K and N440K/E484K substitutions, were generated using site-directed mutagenesis kit according to the manufacturer’s instructions (New England Biolabs (NEB), E0554S). All constructs were confirmed by Sanger sequencing and used to express soluble proteins by transiently transfecting Expi293F cells (GIBCO). Supernatants were harvested after four days, and RBD proteins were purified by nickel affinity chromatography. S 6P proteins were purified by nickel affinity following with size-exclusion chromatography. Peak fractions from size-exclusion chromatography were identified by native gel electrophoresis, and peak fractions corresponding to monomeric RBDs or spike trimers were pooled and stored at 4°C.

### SARS-CoV-2 pseudotyped reporter virus

A plasmid expressing SARS-CoV-2 spike in the context of pSARS-CoV-2-S _Δ19_ (Wuhan-Hu-1) has been described^6^. Two plasmids containing C135/C144 antibody escape mutations were constructed based on pSARS-CoV-2-S_Δ19_ by overlap extension PCR-mediated mutagenesis and Gibson assembly. Specifically, the substitutions introduced were: R346S/Q493K and R346S/N440K/E484K. Those substitutions were incorporated into a spike protein that also includes the R683G substitution, which disrupts the furin cleavage site and increases particle infectivity. Neutralizing activity against mutant pseudoviruses were compared to a wildtype (WT) SARS-CoV-2 spike sequence (NC_045512), carrying R683G where appropriate, as indicated. SARS-CoV-2 pseudotyped particles were generated as previously described^5, 31^. Briefly, 293T cells were transfected with pNL4-3ΔEnv-nanoluc and pSARS-CoV-2-S_Δ19_, particles were harvested 48 hours post-transfection, filtered and stored at −80°C.

### Pseudotyped virus neutralization assay

Fivefold serially diluted pre-pandemic negative control plasma from healthy donors (technical negative controls, data not shown), plasma from vaccinated mAb recipients and mRNA vaccinated controls or monoclonal antibodies were incubated with SARS-CoV-2 pseudotyped virus for 1 hour at 37 °C. The mixture was subsequently incubated with 293T_Ace2_ cells^5^ (for all monoclonal antibody WT neutralization assays) or HT1080Ace2 cl14 cells^32^ (for all plasma neutralization assays) for 48 hours after which cells were washed with PBS and lysed with Luciferase Cell Culture Lysis 5× reagent (Promega). Nanoluc Luciferase activity in lysates was measured using the Nano-Glo Luciferase Assay System (Promega) with the ClarioStar Multimode reader (BMG). The relative luminescence units were normalized to those derived from cells infected with SARS-CoV-2 pseudotyped virus in the absence of plasma or monoclonal antibodies. The half-maximal neutralization titers for plasma (NT_50_) or half-maximal and 90% inhibitory concentrations for monoclonal antibodies (IC_50_ and IC_90_) were determined using four-parameter nonlinear regression (least squares regression method without weighting; constraints: top=1, bottom=0) (GraphPad Prism).

### Biotinylation of viral protein for use in flow cytometry and biolayer interferometry

Purified and Avi-tagged SARS-CoV-2 Wuhan-Hu-1 RBD or S were biotinylated using the Biotin-Protein Ligase-BIRA kit according to manufacturer’s instructions (Avidity) as described before^5^. Ovalbumin (Sigma, A5503-1G) was biotinylated using the EZ-Link Sulfo-NHS-LC-Biotinylation kit according to the manufacturer’s instructions (Thermo Scientific). Biotinylated ovalbumin was conjugated to streptavidin-BV711 for single-cell sorts (BD biosciences, 563262) or to streptavidin-BB515 for phenotyping panel (BD biosciences, 564453). RBD was conjugated to streptavidin-PE (BD Biosciences, 554061) and streptavidin-AF647 (Biolegend, 405237)^5, 11^.

### Flow cytometry and single-cell sorting

Single-cell sorting by flow cytometry was described previously^5^. Briefly, peripheral blood mononuclear cells were enriched for B cells by negative selection using a pan-B-cell isolation kit according to the manufacturer’s instructions (Miltenyi Biotec, 130-101-638). The enriched B cells were incubated in Flourescence-Activated Cell-sorting (FACS) buffer (1× PBS, 2% FCS, 1 mM ethylenediaminetetraacetic acid (EDTA)) with the following anti-human antibodies (all at 1:200 dilution): anti-CD20-PECy7 (BD Biosciences, 335793), anti-CD3-APC-eFluro 780 (Invitrogen, 47-0037-41), anti-CD8-APC-eFluor 780 (Invitrogen, 47-0086-42), anti-CD16-APC-eFluor 780 (Invitrogen, 47-0168-41), anti-CD14-APC-eFluor 780 (Invitrogen, 47-0149-42), as well as Zombie NIR (BioLegend, 423105) and fluorophore-labeled RBD and ovalbumin (Ova) for 30 min on ice. AccuCheck Counting Beads (Life Technologies, PCB100) were added as indicated to each sample according to manufacturer’s instructions. Single CD3-CD8-CD14-CD16−CD20+Ova−RBD-PE+RBD-AF647+ B cells were sorted into individual wells of 96-well plates containing 4 μl of lysis buffer (0.5× PBS, 10 mM Dithiothreitol (DTT), 3,000 units/ml RNasin Ribonuclease Inhibitors (Promega, N2615) per well using a FACS Aria III and FACSDiva software (Becton Dickinson) for acquisition and FlowJo for analysis. The sorted cells were frozen on dry ice, and then stored at −80 °C for subsequent RNA reverse transcription. For B cell isotype analysis by flow-cytometry, in addition to above antibodies, B cells were also stained with following anti-human antibodies (all at 1:200 dilution): anti-CD19-BV605 (Biolegend, 302244), anti-IgG-PECF594 (BD, 562538), anti-IgM-AF700 (Biolegend, 314538), and anti-CD38-BV421 (Biolegend, 303526).

### Antibody sequencing, cloning and expression

Antibodies were identified and sequenced as described previously^5, 33^. In brief, RNA from single cells was reverse-transcribed (SuperScript III Reverse Transcriptase, Invitrogen, 18080-044) and the cDNA was stored at −20 °C or used for subsequent amplification of the variable IGH, IGL and IGK genes by nested PCR and Sanger sequencing. Sequence analysis was performed using MacVector and Geneious Prime (version 2022.1.1). Amplicons from the first PCR reaction were used as templates for sequence- and ligation-independent cloning into antibody expression vectors. Recombinant monoclonal antibodies were produced and purified as previously described^5^.

### Biolayer interferometry

Biolayer interferometry assays were performed as previously described^5^, with minor modifications as below. Briefly, we used the Octet Red instrument (ForteBio) at 30 °C with shaking at 1,000 r.p.m. Monomeric affinities of anti-SARS-CoV-2 RBD IgG binding were derived by subtracting the signal obtained from traces performed with IgGs in the absence of WT RBD. Kinetic analysis using protein A biosensor (ForteBio, 18-5010) was performed as follows: (1) baseline: immersion for 60 s in buffer; (2) loading: immersion for 200 s in a solution with IgGs at 10 μg ml–1; (3) baseline: immersion for 200 s in buffer; (4) association: immersion for 300 s in solution with WT RBD at three different concentrations ranging from 200 to 5 μg ml–1; (5) dissociation: immersion for 600 s in buffer. Curve fitting was performed using a fast 1:1 binding model and the data analysis software from ForteBio. Mean equilibrium dissociation constants (Kd) were determined by averaging all binding curves that matched the theoretical fit with an R2 value ≥0.8. To establish binding of low-affinity antibodies to multimerized antigen, 6P-stabilized and biotinylated S trimers were incubated with recombinant Streptavidin (ACROBiosystems, STN-N5116) for 30 min at RT, resulting in up to12 RBD-binding moieties per molecule and assayed on the Octet Red instrument (ForteBio) as above, with the following modification: Association step (4) was performed with the S-multimer at 430 μg ml–1. Epitope mapping assays were performed with protein A biosensor (ForteBio 18-5010), following the manufacturer’s protocol “classical sandwich assay” as follows: (1) Sensor check: sensors immersed 30 sec in buffer alone (buffer ForteBio 18-1105), (2) Capture 1st Ab: sensors immersed 10 min with Ab1 at 10 µg/mL, (3) Baseline: sensors immersed 30 sec in buffer alone, (4) Blocking: sensors immersed 5 min with IgG isotype control (3BNC117) at 20 µg/mL. (5) Baseline: sensors immersed 30 sec in buffer alone, (6) Antigen association: sensors immersed 5 min with RBD at 20 µg/mL. (7) Baseline: sensors immersed 30 sec in buffer alone. (8) Association Ab2: sensors immersed 5 min with Ab2 at 10 µg/mL. Curve fitting was performed using the Fortebio Octet Data analysis software (ForteBio).

### Computational analyses of antibody sequences

Antibody sequences were trimmed based on quality and annotated using Igblastn v.1.14. with IMGT domain delineation system. Annotation was performed systematically using Change-O toolkit v.0.4.540^34^. Clonality of heavy and light chain was determined using DefineClones.py implemented by Change-O v0.4.5^34^. The script calculates the Hamming distance between each sequence in the data set and its nearest neighbor. Distances are subsequently normalized and to account for differences in junction sequence length, and clonality is determined based on a cut-off threshold of 0.15. Heavy and light chains derived from the same cell were subsequently paired, and clonotypes were assigned based on their V and J genes using in-house R and Perl scripts. All scripts and the data used to process antibody sequences are publicly available on GitHub (https://github.com/stratust/igpipeline/tree/igpipeline2_timepoint_v2).

The frequency distributions of human V genes in anti-SARS-CoV-2 antibodies from this study was compared to 131,284,220 IgH and IgL sequences generated by Soto et al.^35^ and downloaded from cAb-Rep^36^, a database of human shared BCR clonotypes available at https://cab-rep.c2b2.columbia.edu/. Based on the 108 distinct V genes that make up the 417 analyzed sequences from the Ig repertoire of the individuals described in this study (353 sequences isolated form 5 monoclonal antibody recipients and 65 IgM sequences isolated from 9 vaccinated control individuals (for IgG sequences isolated from controls see ^10, 11^), we selected the IgH and IgL sequences from the database that are partially coded by the same V genes and counted them according to the constant region. The frequencies shown in Extended Data Fig. 4 are relative to the source and isotype analyzed. We used the two-sided binomial test to check whether the number of sequences belonging to a specific IGHV or IGLV gene in the repertoire is different according to the frequency of the same IgV gene in the database. Adjusted p-values were calculated using the false discovery rate (FDR) correction. Significant differences are denoted with stars.

Nucleotide somatic mutations and Complementarity-Determining Region (CDR3) length were determined using in-house R and Perl scripts. For quantification of somatic mutations, *IGHV* and *IGLV* nucleotide sequences were aligned against their closest germlines using Igblastn and the number of differences were considered nucleotide mutations.

## Data Availability

All data produced in the present study are available upon reasonable request to the authors.

## Data presentation

Figures arranged in Adobe Illustrator 2022.

## Data availability statement

Data are provided in Supplementary Tables 1-4. The raw sequencing data and computer scripts associated with Figure 2 and Ext. Data Fig. 3 have been deposited at Github (https://github.com/stratust/igpipeline/tree/igpipeline2_timepoint_v2). This study also uses data from “A Public Database of Memory and Naive B-Cell Receptor Sequences” (https://doi.org/10.5061/dryad.35ks2), PDB (6VYB and 6NB6), cAb-Rep (https://cab-rep.c2b2.columbia.edu/), Sequence Read Archive (accession SRP010970), and from “High frequency of shared clonotypes in human B cell receptor repertoires” (https://doi.org/10.1038/s41586-019-0934-8).

## Code availability statement

Computer code to process the antibody sequences is available at GitHub (https://github.com/stratust/igpipeline/tree/igpipeline2_timepoint_v2).

## Acknowledgements

We thank all study participants who devoted their time to our research; The Rockefeller University Hospital nursing staff and Clinical Research Support Office and nursing staff; C. M. Rice, and all members of the M.C.N. laboratory for helpful discussions; M. Jankovic and G. Scrivanti for laboratory support; K. Gordon for cell sorting. This work was supported by NIH grants P01-AI138398-S1 and 2U19-AI111825 to M.C.N., R37-AI64003 to P.D.B. and R01-AI78788 to T.H., and 3UM1AI126620-5S1 to M. Caskey. D.S.-B. is supported in part by the National Center for Advancing Translational Sciences (NIH Clinical and Translational Science Award program, grant UL1-TR001866) and the Shapiro–Silverberg Fund for the Advancement of Translational Research. F.M. was supported by the Bulgari Women & Science Fellowship in COVID-19 Research. P.D.B. and M.C.N. are Howard Hughes Medical Institute (HHMI) Investigators. This article is subject to HHMI’s Open Access to Publications policy. HHMI lab heads have previously granted a nonexclusive CC BY 4.0 license to the public and a sublicensable license to HHMI in their research articles. Pursuant to those licenses, the author-accepted manuscript of this article can be made freely available under a CC BY 4.0 license immediately upon publication.

## Author Contributions

D.S.-B. and M.C.N. conceptualized the study. D.S.-B., F.M., Z.W., A.C., P.D.B., T.H., and M.C.N. conceived, designed and analyzed experiments. D.S.-B., C.G., and M. Caskey designed clinical protocols. D.S.-B., Z.W., F.M., A.C., R.R., M. Canis, J.DaSilva., F.S. and K.Y. carried out experiments. B.J. and A.G. produced antibodies. M.T., K.G.M., I.S., J.Dizon, C.G. and M. Caskey recruited participants, executed clinical protocols, and processed samples. T.Y.O. and V.R. performed bioinformatic analysis. D.S.-B. and M.C.N. wrote the manuscript with input from all co-authors.

## Extended Data Figures

**Extended Data Fig. 1:**
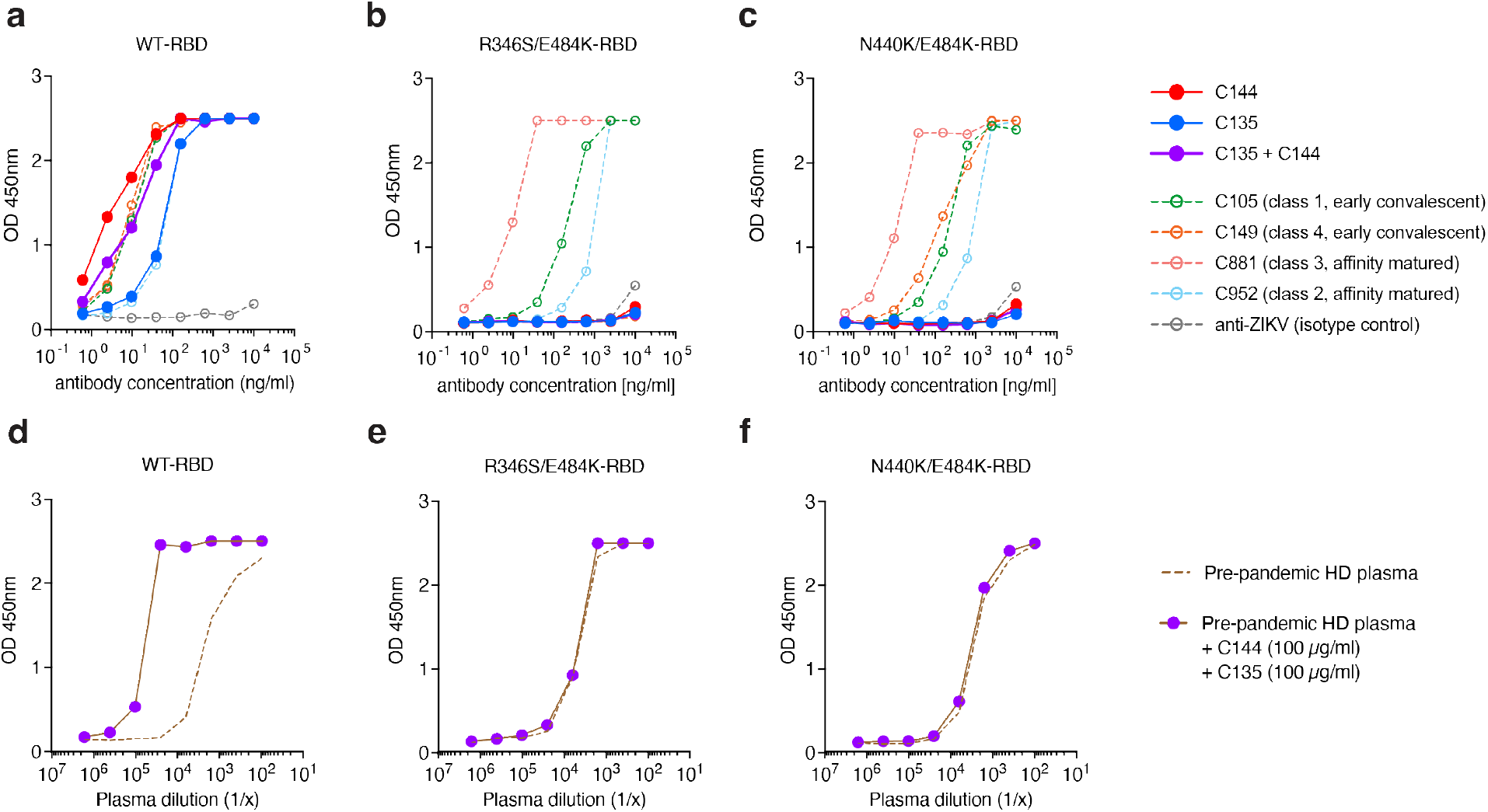
Mutant RBDs selectively abrogate binding by C135 and C144. Monoclonal antibody binding to mutant forms of RBD. **a-c,** Graphs show concentration-dependent antibody binding to (**a**) WT, (**b**) R436S/E484K, and (**c**) N440K/E484K RBDs by C144, C135, and Class 1 (C105), Class 2 (C952), Class 3 (C881), and Class 4 (C149)^5, 6, 8, 23^. **d-f,** Graphs show concentration dependent pre-pandemic healthy donor plasma binding to (**d**) WT, (**e**) R436S/E484K, and (**e**) N440K/E484K RBDs in the presence (purple) or absence (dotted lines) of 100μg/ml of C135 and C144. Addition of C144 and C135 to plasma increases the binding activity of plasma against the WT but not the 2 mutant RBDs.

**Extended Data Fig. 2:**
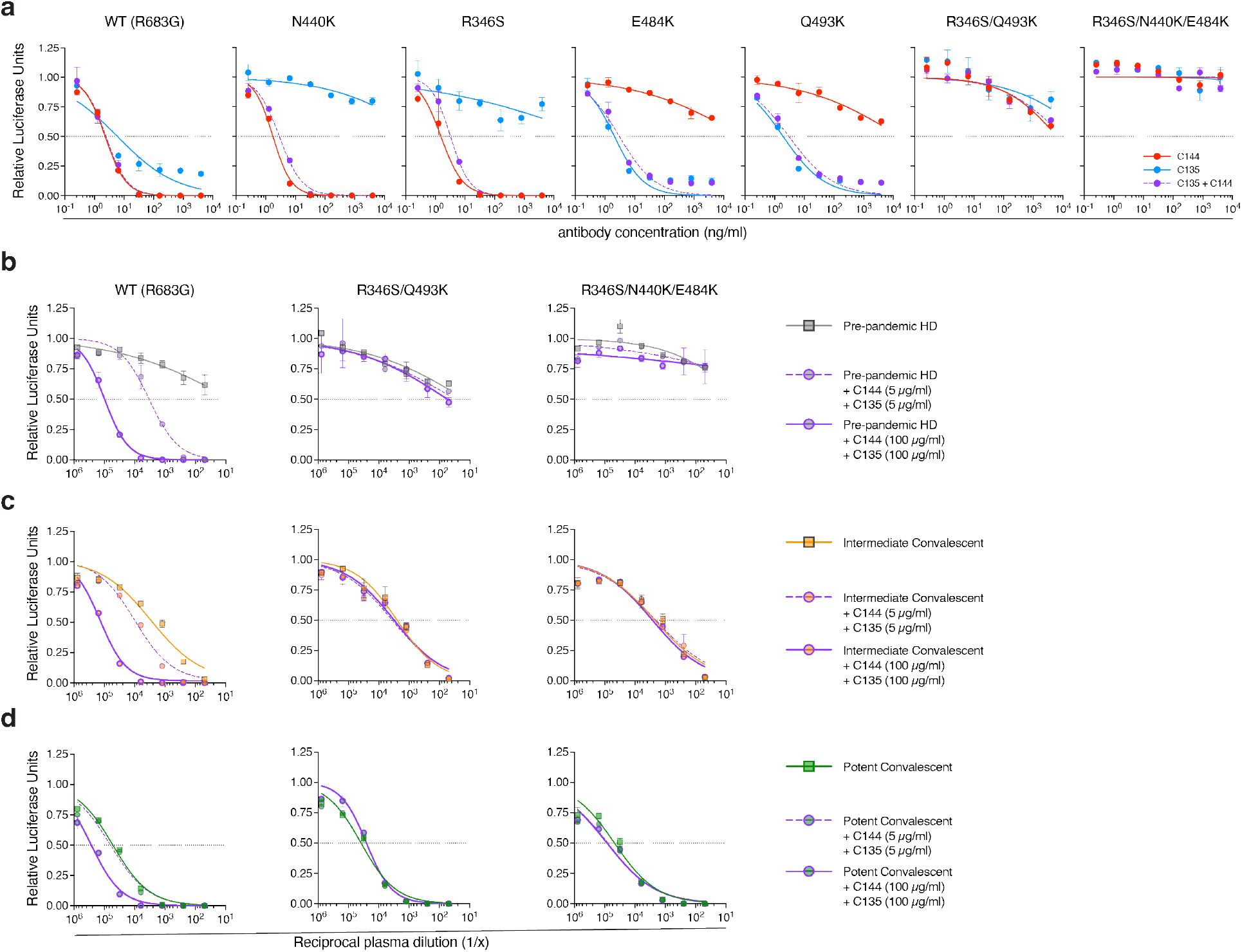
SARS-CoV-2 R346S/Q493K and R346S/N440K/E484K pseudotype virus neutralization by C135-LS and C144-LS. **a,** Graphs show concentration-dependent neutralization curves for SARS-CoV-2 pseudoviruses by monoclonal antibodies. C144 (red), C135 (blue), and their equimolar combination (purple). **b,** Pre-pandemic plasma (squares) neutralization of WT or R346S/Q493K or R346S/N440K/E484K pseudoviruses in the absence or presence of 5 (purple dashed circles) or 100μg/ml (purple solid circles) of C135 and C144^14^. **c-d,** As in (**b**) but for convalescent plasma with intermediate (**c**, COV157) ^12^ or strong (**d**, COV31)^12^ neutralizing activity. The horizontal lines in all panels indicate half-maximal neutralization.

**Extended Data Fig. 3:**
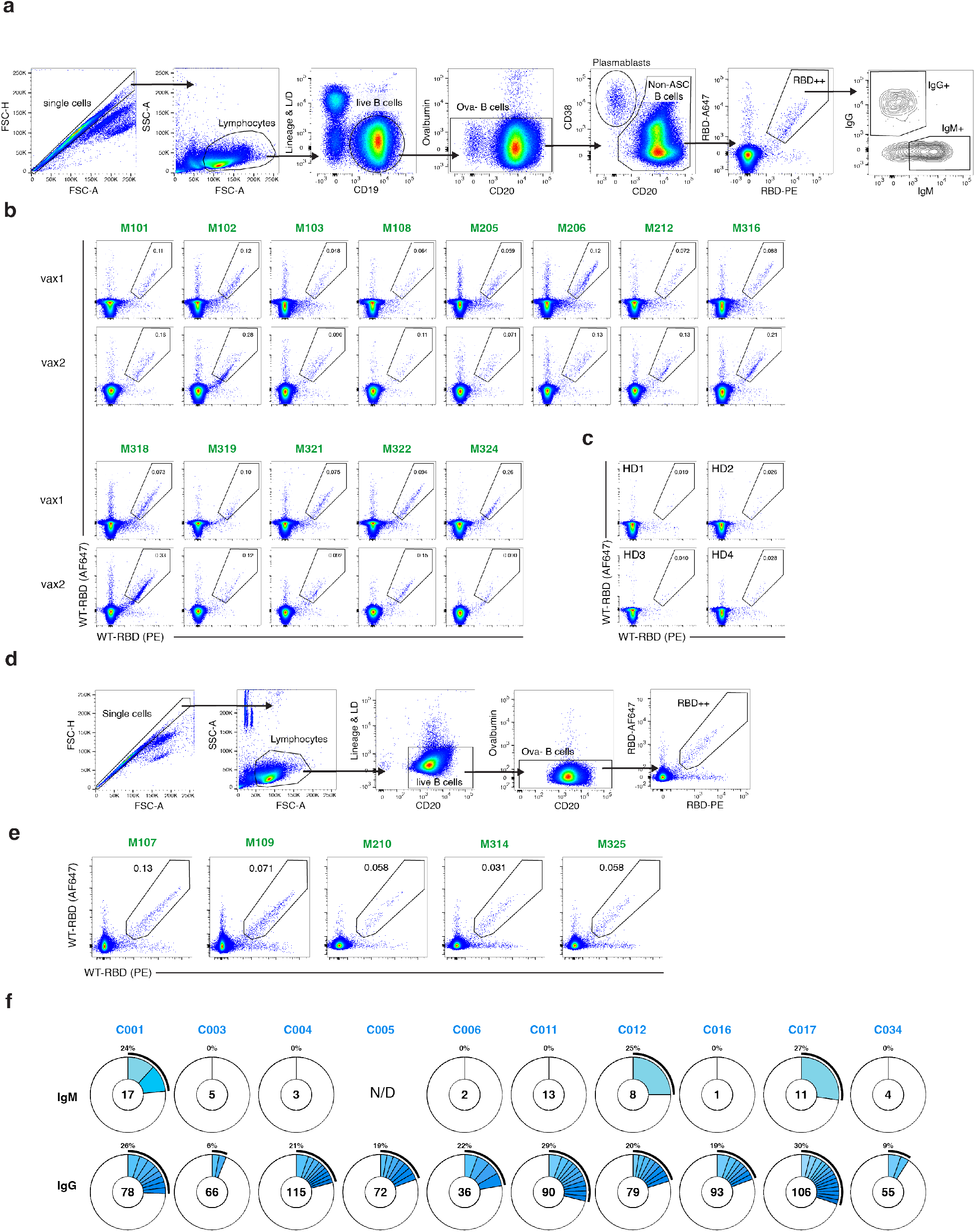
Flow cytometry, single-cell sorting and BCR sequencing. **a,** Gating strategy for flow-cytometry phenotyping. Gating was on single lymphocytes that were CD19^+^ and CD20^+^, and CD3^-^ CD8^-^ CD16^-^ Ova^-^ without uptake of live-dead dye (L/D). Antigen-specific cells were those with dual binding to Wuhan-Hu1 RBD-PE and RBD-AF647. Anti-IgG, -IgM were used to phenotype dual RBD-labelled B cells. **b,c,** Representative flow cytometry plots of Wuhan Hu-1 RBD-binding memory B cells from 13 mAb recipients after one and two doses of vaccination (**b**) and pre-pandemic health donors (**c**) serving as negative controls. Numbers in RBD-gate denote percentage of RBD dual-labelled cells of parent gate (see **a**). Corresponding flow-cytometry plots and gating strategy for vaccinated controls can be found in ^10^ **d,** Gating strategy for single-cell sorting of RBD-specific memory B cells. Dual-labelled (RBD-PE^+^/-AF647^+^) CD20^+^ CD3^-^ CD8^-^ CD16^-^ Ova^-^ cells were sorted. **e,** Representative flow cytometry plots show RBD-binding cells that were sorted from 5 mAb recipients (Fig. 2g). **f,** Pie charts show the distribution of antibody sequences derived from cells isolated from 10 vaccinated control individual after vax2 (Fig 2h). The upper panel shows IgM, and the lower panel depicts IgG sequences^10, 11^. The number in the inner circle indicates the number of sequences analyzed for the individual denoted above the circle. Slices colored in shades of blue indicate cells that are clonally expanded (same IGHV and IGLV genes, with highly similar CDR3s). Pie slice size is proportional to the number of clonally related sequences. The black outline and % value indicate the frequency of clonally expanded sequences detected within an individual. White pie areas indicate the proportion of sequences isolated only once. For C005, there were no IgM transcripts amplified at the timepoint assayed.

**Extended Data Fig. 4:**
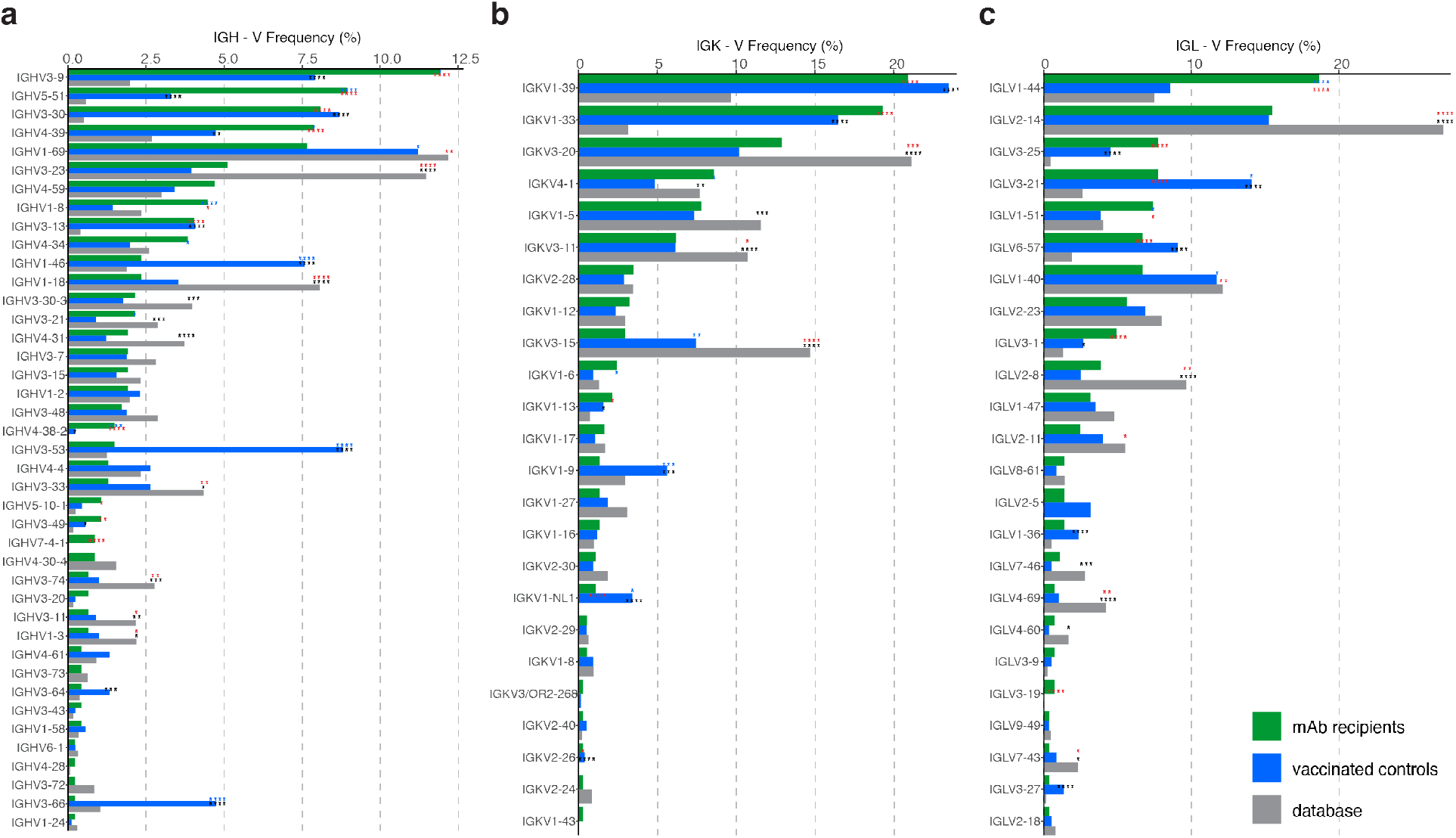
Frequency distribution of human V genes. **a-c,** Comparison of the frequency distribution of V gene usage for the IgH and IgL among antibodies isolated from mRNA-vaccinated mAb recipients (this study) and controls^10, 11^, after vax2, and from database of shared clonotypes of human antibodies from Soto et al^35^. Graphs show relative abundance of human IGHV (**a**), IGKV (**b**), and IGLV (**c**) genes within the human V gene database (in grey, Sequence Read Archive accession SRP010970), antibodies isolated from mAb recipients (in green) or vaccinated controls (in blue). Colors of stars indicate levels of statistical significance for the following frequency comparisons: black – vaccinated controls vs. database; red – mAb recipients vs. database; blue – mAb recipients vs. vaccinated controls.

**Extended Data Fig. 5:**
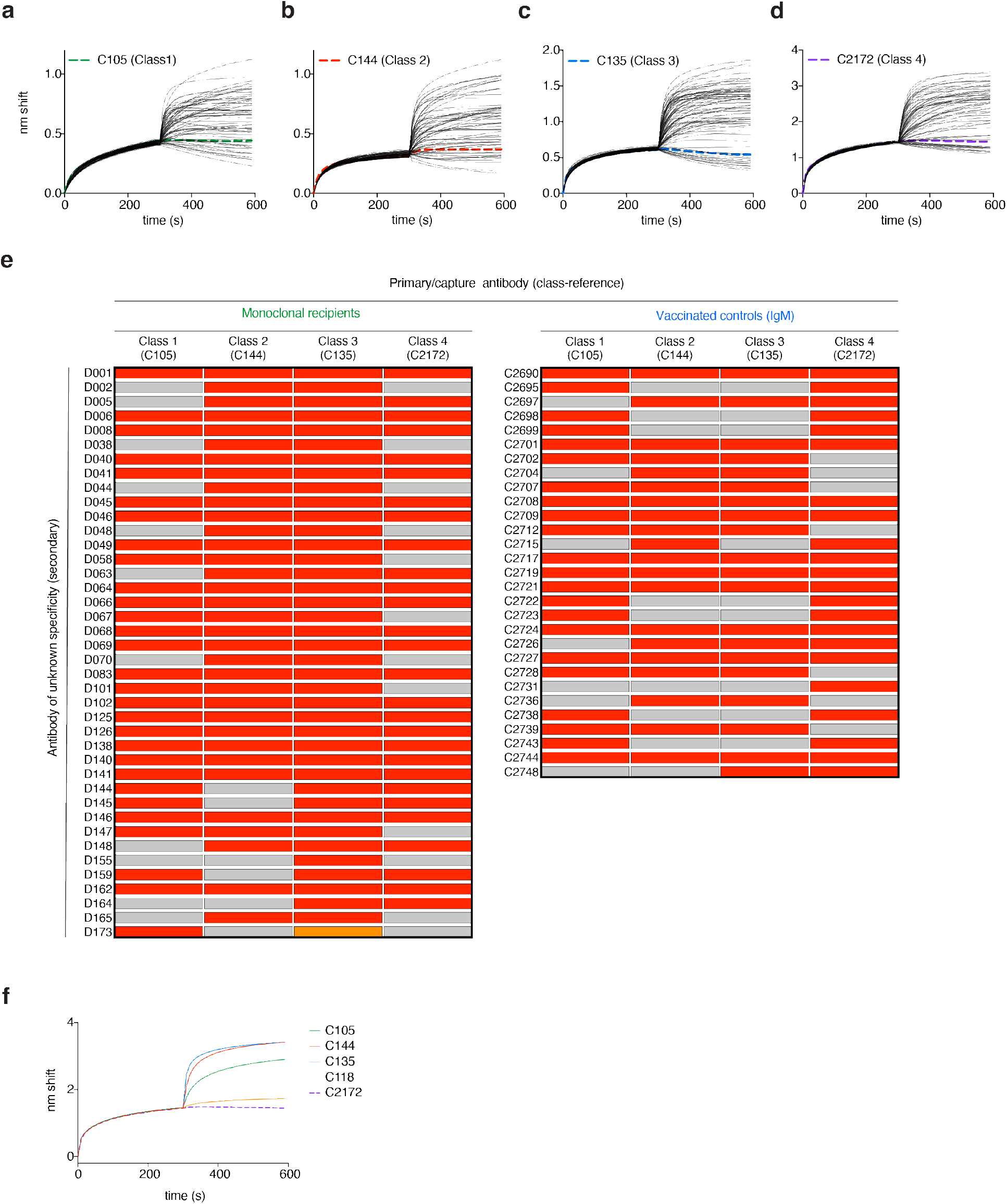
Competition BLI. **a-d,** BLI traces of antibodies assayed for competition with class-reference antibodies. Traces show initial association curve (antigen capture phase of the primary antibody) and subsequent addition of secondary antibodies of unknown class. Thin solid black lines represent antibodies isolated from mAb recipients or vaccinated controls. Thick dashed lines are self-competition traces of C105 (green in **a**), C144 (red in **b**), C135 (blue in **c**) and C2172 (purple in **d**) for classes 1-4, respectively. **e,** Heat-map of relative inhibition of secondary antibody binding to the preformed capture antibody-RBD complexes (grey=no binding, red=unimpaired binding, orange=indeterminate). The left panel shows antibodies from mAb recipients, while the right panel shows IgM antibodies from vaccinated controls isolated in this study (both after vax2). Details on IgG antibodies isolated from vaccinated controls can be found in ^10, 11^. **f,** BLI traces defining C2172 as Class 4. C2172 is the primary/capture antibody (in dashed purple). The addition of known class-defining antibodies C105 (in green, Class 1^8^), C144 (in red, Class 2^8^), C135 (in blue, Class 3^8^), and C118 (in orange, Class 1/4^24^) establish C2172 as a bona fide Class 4 antibody.

